# Spatiotemporal Trends in Bed Bug Metrics, New York City

**DOI:** 10.1101/2020.06.30.20143669

**Authors:** Kathryn P. Hacker, Andrew J. Greenlee, Alison L. Hill, Daniel Schneider, Michael Z. Levy

## Abstract

Bed bug outbreaks pose a major challenge in urban environments and cause significant strain on public resources. Few studies have systematically analyzed the epidemic or the potential effects of policies to combat bed bugs. Here we use three sources of administrative data to characterize the spatial-temporal trends of bed bug inquiries, complaints, and reports in New York City. From 2014–2020, Bed bug complaints have significantly decreased (p < 0.01), the absolute number of complaints per month dropping by half (875 average complaints per month to 440 average complaints per month); conversely cockroach-specific complaints increased over the same period. Despite the decrease of bed bug complaints, areas with reported high bed bug infestation tend to remain infested, highlighting the persistence of these pests. There are limitations to the datasets; still the evidence available suggests that interventions employed by New York City residents and lawmakers are stemming the bed bug epidemic and may serve as a model for other large cities.

## Introduction

Bed bugs (*Cimex lectularius*) have reemerged as a substantial public health and economic issue, particularly in dense urban environments ^1–4^. While bed bugs were largely controlled after the Second World War, their populations have resurged since. The resurgence of bed bug populations is likely due to a combination of many factors, among these insecticide resistance ^5–7^, increased mobility ^7–9^, and exchange of used furniture ^10–12^. By the early 1990s, bed bugs were again documented globally as an arthropod pest of public health importance ^1–3,6,13,14^.

The overall prevalence of bed bug infestation in major US cities is high, though rarely systematically measured, and has attracted the attention of the media and policy entities ^15,Supplemental Citation 13, 15^. Trends in bed bug resurgence and control effectiveness are poorly understood. In 2014, New York City established a reporting system for bed bug infestation through the city’s 311 database, a dedicated phone and online system to access NYC services and information ^SupplementalCitation 7,15^. Additionally, in 2014, the New York City department of Health and Human Services collected data on probable bed bug infestations, estimating a prevalence of 5.1% of households citywide, with some areas (defined by New York United Hospital Fund Regions) reporting up to 12% of households infested ^16^. Other cities across the United States have experienced high levels of infestation and have responded with various policy and public health interventions ^15,17^.

The public health burden of bed bug infestations is substantial. Not only are residents of infested dwellings subject to physical symptoms such as painful bites, rashes, sleep loss, and allergic reactions, but some also suffer immense psychological and emotional distress ^5,13,18–20^. Residents of infested dwellings report increased anxiety and depression, linked both to the physicality of the infestation as well as the incurred social stigma ^20,21^. The health burden on the homebound and elderly is particularly relevant, as home healthcare personnel and social workers without adequate training can be reticent, or refuse to, enter infested areas ^22,23^. Additionally, due to the expense and difficulty of effective extermination, poisoning, property damage, and exposure to inexpertly applied insecticides has occurred ^23,24^. Bed bugs are competent hosts for *Trypanosoma cruzi* and *Bartonella quintana*, the etiologic agents of Chagas disease and trench fever respectively ^5,25,26^. Whether bed bugs are, or could become, epidemiologically relevant in the transmission of these agents remains unclear.

To combat this public health crisis, New York City has instituted two bed bug disclosure policies. In 2010, New York passed its first ordinance that required landlords to report bed bug infestations occurring in the previous year to residents and prospective residents ^Supplemental Citation 1^. The city passed a second disclosure ordinance in 2017 requiring landlords to report annually all units infested or treated for bed bug infestation, and to notify all residents in the building, rather than only current or prospective tenets of a given unit ^Supplemental Citation 2^.

Making use of data resulting from 311 and other reporting systems, we assess the spatial-temporal trends in bed bug complaints and inquiries made by New York City residents and building owners. In addition to exploring these trends, we question whether the policy-driven approaches to managing bed bug infestations have resulted in a decrease in the rate of complaints over time. We also question whether these policy approaches have had differential impacts across New York City’s boroughs.

## Methods

### Databases

We examine three databases archived by New York City: bed bug inquiries registered by the city’s 311 non-emergency reporting system, official bed bug complaints made to the city’s Department of Housing Preservation and Development, and building owner reported bed bug infestations reported to the city’s Department of Housing Preservation and Development. The specifics of each database (attributes, time scales, and geographic information) are summarized in Supplemental Table 1 and described in detail below.

### NYC Open Data and NYC 311: 311 inquiries and 311 bed bug specific requests

Since 2009, New York City’s Open Data Portal has maintained an online database of information collected by the city government. One of the largest datasets is the 311 service, a designated system for non-emergency information and reporting, which allows individuals, organizations, and businesses to access New York City’s government services and information ^Supplemental Citation 5^. We accessed all 311 data inquiries from 2010–2019 focusing on 311 inquiries specific to bed bugs (all bed bug related inquiries as well as official complaints).

### NYC housing maintenance code complaints: official bed bug complaints

Under the Housing and Maintenance Code tenants have the right to a bed bug free environment ^Supplementary Citation 1^. Specifically, in the Housing and Maintenance Code, Subchapter 2, Article 4 names bed bugs as a Class B violation, meaning that the landlord is legally obligated to eradicate (*sic*) the problem within 30 days ^Supplementary Citation 1^. These official complaints were made publicly available in 2014 and are updated continuously. We compiled all Housing Maintenance Code Complaints from 2014–2019 that were the result of a potential bed bug complaint, and, as a point of comparison, also compiled cockroach infestation complaints from the same data.

### Building owner reports of bed bug infestation

The newest bed bug disclosure law requires property owners of multiple dwellings (buildings with 3+ residential units) to report annually the number of units infested with bed bugs or that were treated for bed bugs ^Supplemental Citation 2^. While this data is required to be accessible to the public, currently New York City does not have this data published through NYC Open Data. We obtained bed bug infestations reported by property owners for 2018 through a Freedom of Information Request. Data were reported at the building level and included information on the total number of residential units, the number that had experienced a bed bug infestation, the number of infestations treated, and the number of units re-infested.

### Statistical Analyses

#### Assessing the geographical distribution of bed bug infestation in New York City

To assess the geographical distribution of bed bug complaints we calculated the number of bed bug complaints per Neighborhood Tabulation Area (NTA). NTAs are combinations of whole census-tract level population data with a minimum of 15,000 residents per aggregation ^Supplemental Citation 6^. Unlike census tracts, which are prone to high sampling error, using NTAs as a geographic boundary helps to standardize areas by population while providing a more statistically reliable estimate of population ^Supplemental Citation 6^. We divided the total number of bed bug complaints by the estimated population for each year per 100,000. We mapped this information using QGIS software ^Supplemental Citation 11^.

#### Modeling the Temporal Patterns of Housing Maintenance Code Complaints for Bed Bugs and Cockroaches

We modeled the number of complaints per month as a function of time using linear regression incorporating a sinusoidal curve with linearly decreasing amplitude over time. The inclusion of this curve helped account for the high degree of seasonality observed. Since we observed an overall decrease in total number of 311 inquires over time, we standardized the total number of reported infestations to the total number official complaints made through 311. We repeated this analysis for each of the New York City boroughs, standardizing the number of complaints per borough using population estimates obtained by the American Community Survey ^Supplemental Citation 7^.

#### Spatial-Temporal Model

To assess the spatial-temporal dynamics of bed bug complaints, we first calculated the ratio between the observed and expected counts (Standard Incidence Ration SIR) of bed bug complaints per census tract ^27^. We then used a Bayesian Hierarchal modeling approach using INLA, specifically a non-parametric dynamic space-time model, that assessed the relative risk of an NTA area reporting a bed bug complaint ^28^. This approach enabled us to utilize information from neighboring NTA areas and incorporate space-time covariates. This spatial-temporal approach accounts for not only spatial structure, but also temporal and spatial-temporal interactions, and smooths or shrinks extreme values that would potentially result by using SIR values alone^28,29^.

#### Comparing landlord reports of bed bug infestation to 311 resident complaints for 2018

We examined the concordance between the 311-complaint data and recently available data on building owner reports for each NTA using linear regression.

## Results

### Summary of 311 inquiries linked to bed bugs from 2010 - 2019

From 2010–2019 there were a total of 72,701,278 inquiries processed by 311 either online, through the app, or by phone. Since 2010, the number of inquiries processed by 311 has steadily decreased (Fig. 1). We identified 185,289 inquiries specific to bed bugs. These inquiries were processed as 18 specific descriptions which were forwarded to seven different city departments or by the general 311 call center (Supplementary Table 2).

**Fig. 1.**
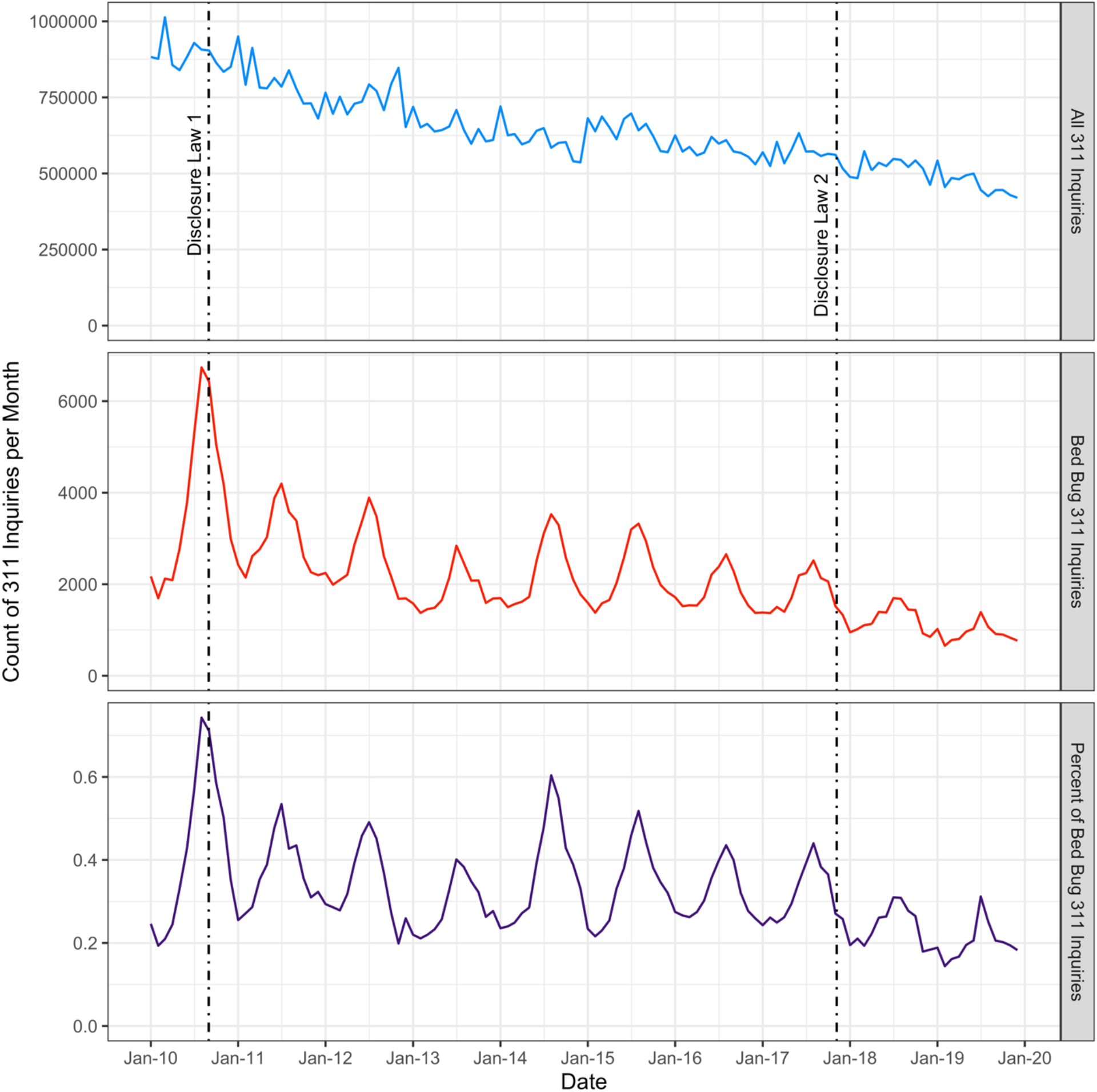
New York City 311 data usage and bed bug related inquiries. New York City 311 inquiries, bed bug related inquiries (which include official bed bug complaints registered to the Department of Housing Preservation, and bed bug related inquiries standardized by the total number of 311 inquiries, from 2010–2019. Bed bug related inquires were extracted using a text search algorithm. Dates of the two bed bug disclosure laws are indicated by a dashed line.

Similar to the general 311 inquiries, bed bug related inquiries decreased from 2010–2019. The largest peak in bed bug related inquiries occurred in August 2010 (n = 6,737). Throughout the time series, bed bug inquiries peaked during late summer (June–August) and decreased from September through April, creating a distinct seasonal pattern.

### Temporal Patterns of Housing Maintenance Code Complaints for Bed Bugs and Cockroaches

Official bed bug complaints followed seasonal patterns (Fig. 2A, Supplemental Table 4). When this harmonic pattern was extracted, the residuals formed a linear decreasing trend from 2014–2019. The decreasing temporal trend was significant, indicating that bed bug complaints significantly decreased from 2014–2019 (Supplemental Table 3). Cockroach complaints followed a similar seasonal pattern to bed bug complaints (Fig. 2B). However, unlike bed bug complaints, we observed a significant positive temporal trend (Supplemental Table 3).

**Fig. 2.**
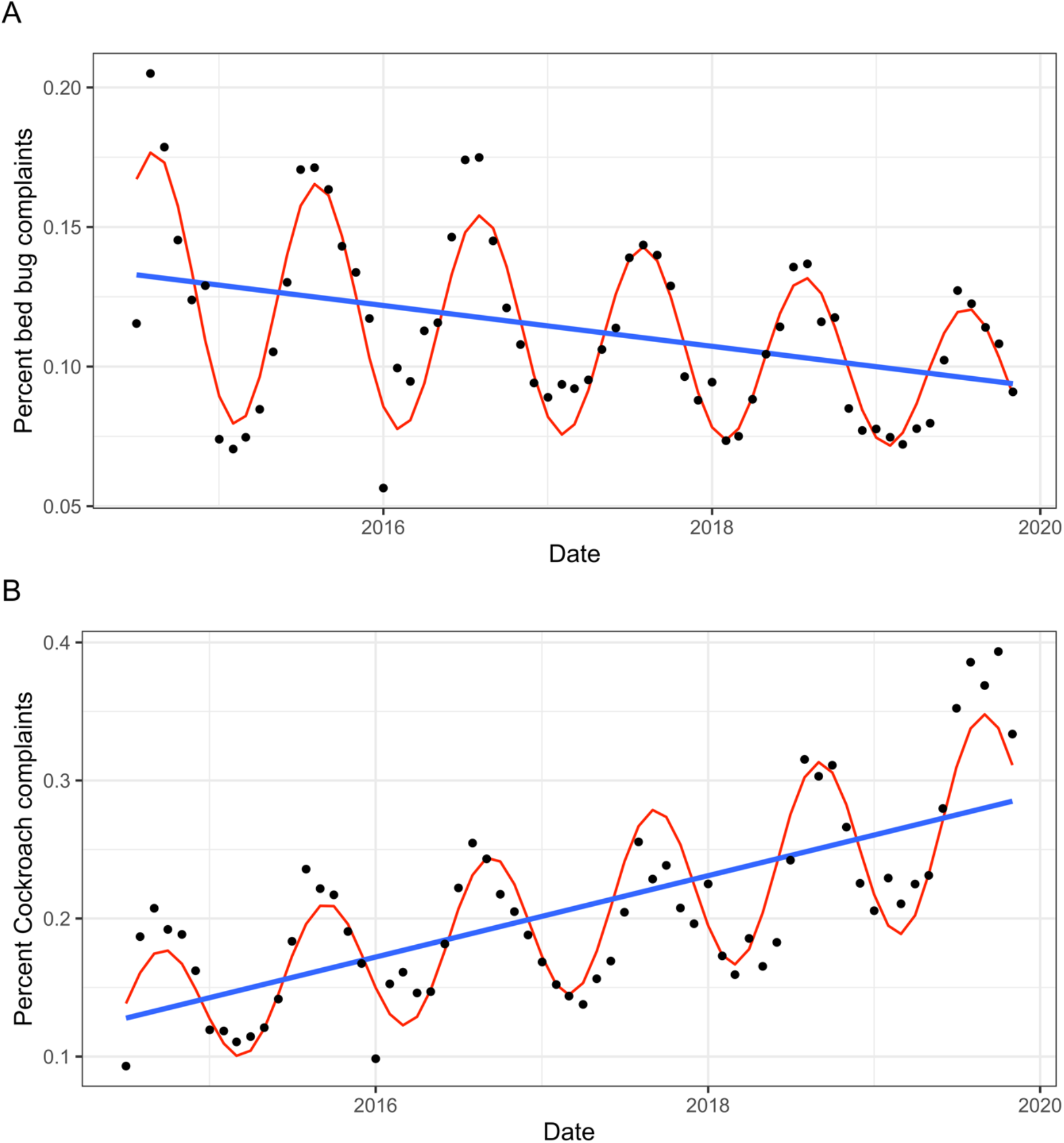
Longitudinal analysis of official bed bug and cockroach complaints registered to HPD. Graphical representation of the results of a linear harmonic model assessing the temporal relationship of official bed bug and cockroach complaints from 2014–2019. (*A*) Percent Bed bug (Bed bug complaints/total number of inquiries) complaints modeled as a linear harmonic model with decreasing amplitude over time. (*B*) Cockroach complaints (cockroach complaints/total number of inquiries) modeled as a linear harmonic model with increasing amplitude over time. (*B*) linear harmonic model assessing official cockroach complaints over time.

Across all the boroughs, bed bug complaints decreased (Fig. 3). Brooklyn had the greatest yearly rate of decrease, followed by the Bronx and Manhattan, Queens and lastly Staten Island (Supplemental Table 3, Supplemental Fig. 1). The seasonal pattern was evident across all the boroughs.

**Fig. 3.**
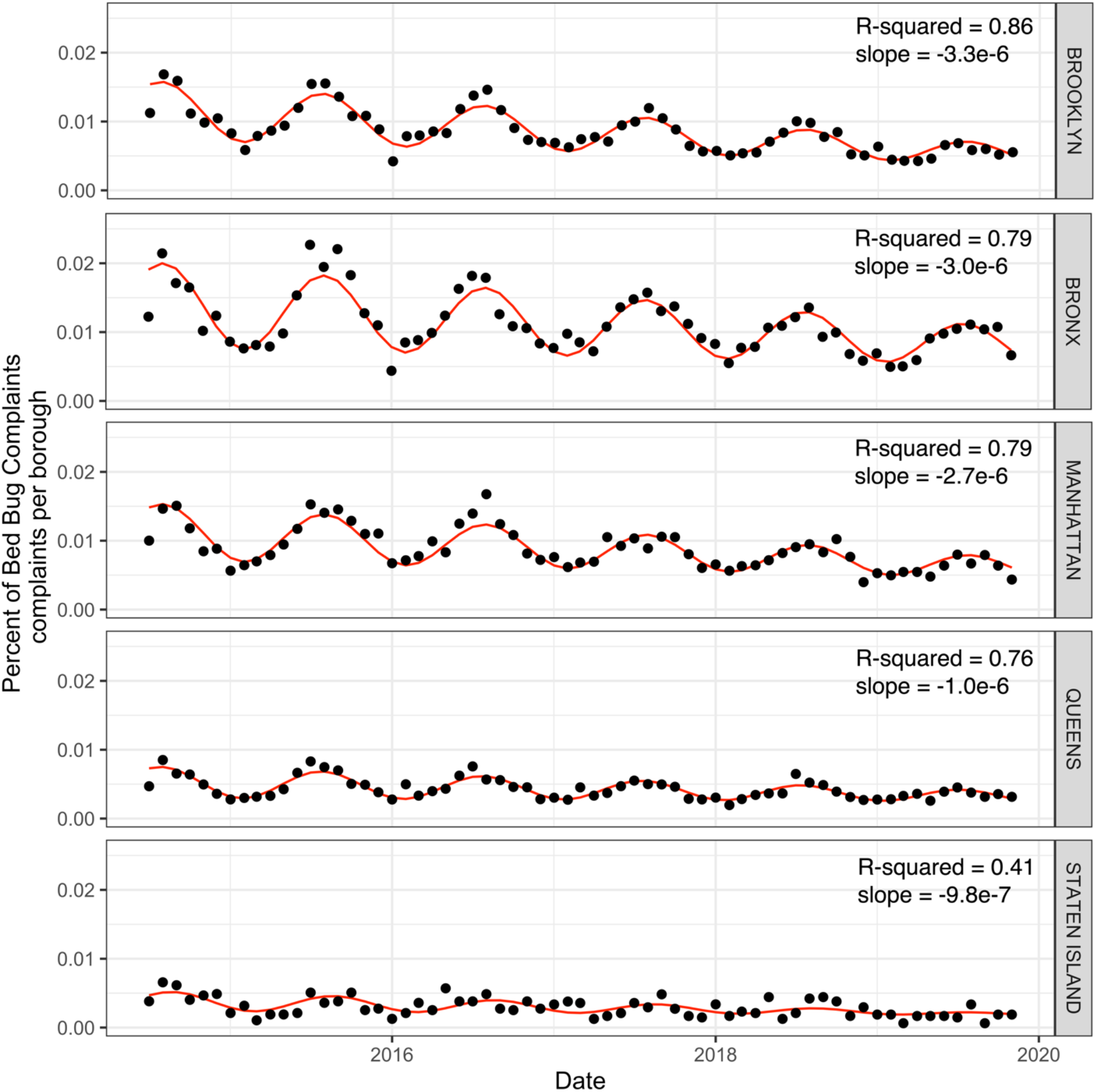
Harmonic linear model assessing the temporal patterns of official bed bug complaints for each of the five NYC boroughs. Graphical representation of model results assessing official bed bug complaints per each of the NYC borough standardized by borough population from 2014– 2019. Model fit assessed by R-squared and slope of the linear residual pattern are reported.

### Spatiotemporal modeling of bed bug complaints in New York City

From 2014–2019, the number of official bed bug complaints processed by HPD were widely distributed throughout the five boroughs (Fig. 4, Supplemental Fig. 1). The Bronx had the greatest proportional number of bed bug complaints followed by Brooklyn, Manhattan, Queens, and Staten Island for the study period (Supplemental Table 4). During 2015, bed bug complaints peaked and then decreased across all boroughs (Supplemental Table 4).

**Fig. 4.**
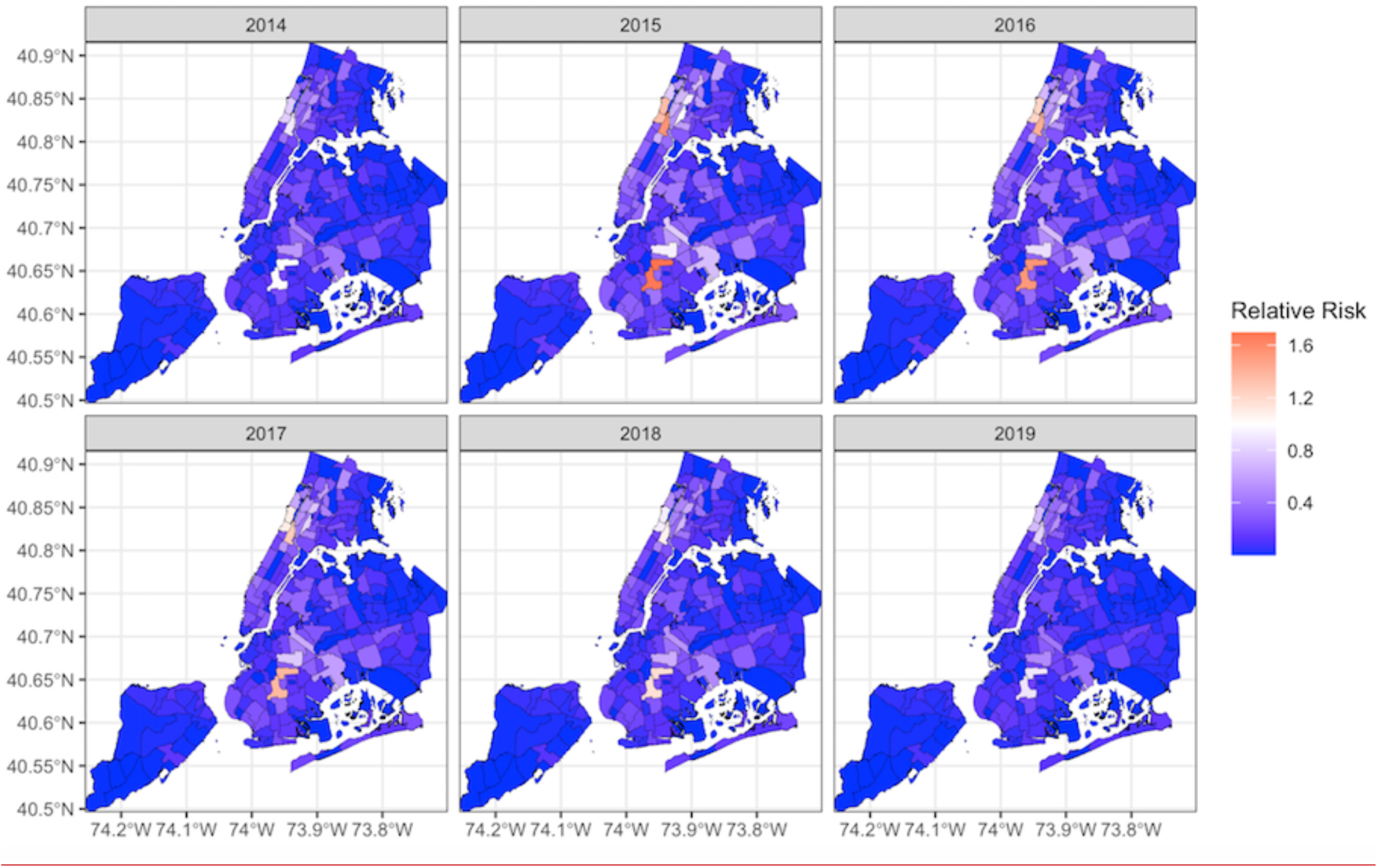
Relative risk of official bed bug complaints per NTA area incorporating a temporally structured fixed effect. The model incorporates neighborhood effects, where the surrounding NTA areas influence the overall relative risk of the central NTA area. This spatiotemporal approach enables us to examine relative risk while adjusting for both space and time.

The non-parametric dynamic space-time model incorporated the addition of neighborhood effects, where the relative risk of neighboring NTA areas influences the relative risk of each NTA area. The relative risk in some NTA areas were influenced by neighborhood effects. Of note, these effects were noticeable in Brooklyn in 2016 and 2017, however neighborhood effects did not contribute significantly to relative risk in the majority of NTA areas. When random spatial, temporal, and spatiotemporal effects were integrated into the model, the geographic distribution and relative intensities of risk did not differ substantially from the originally computed Standard Incidence Ratio (Fig. 4 vs Supplemental Fig. 1). Put differently, the space-time model with neighborhood effects adequately captures the spatiotemporal trends and was not improved with random effects for space or time.

### Comparing landlord reports of bed bug infestation to 311 resident complaints for 2018

When we compared 2018 HPD bed bug complaint data (total number of complaints per NTA) to building owner reported bed bug infestation (number of reports per NTA) we found significant positive agreement (0.92 ± CI 0.81, 1.02, p < 0.001), suggesting that although only a portion of infestations may be officially reported, we expect complaints and reports to follow the same spatiotemporal trends. The correlation was high (R2= 0.60).

## Discussion

During the time periods assessed, both general 311 inquiries about bed bugs (2010–2019) and official bed bug complaints (2014–2019) decreased in New York city. This decrease was significant both when standardized by population and against the totality of 311 inquiries and is directly contrasted to patterns observed in cockroach complaints. These contrasting patterns, as well as strong seasonal patterns, indicate that the decrease in complaints are not an artifact of overall 311 or reporting use. Ultimately, disclosure laws, new approaches in pest management, increased knowledge, and commitment to inspections have likely all contributed to this decrease in bed bug complaints in New York City.

New York City has enacted one of the most comprehensive strategies to combat bed bug infestations among major US cities ^15^. Ordinances assign responsibility for treatment to landlords, and subsequent disclosure of infestations to tenants. The city then commits substantial resources to rapidly respond to all bed bug related complaints, even employing a canine team to help assist in timely inspections ^30^. While our study cannot individually and specifically assess the efficacy of these policies, the decrease in bed bug complaints across all boroughs provides evidence that they are working. A previous modeling study by Xie et al, not only demonstrates that disclosure laws can reduce the spread of bed bug infestations, but also that they can reduce the costs incurred by landlords and tenants ^31^. Strong disclosure laws, like those in New York City, may therefore offer a cost-effective road map for other cities struggling with bed bugs.

Best practices for bed bug management have also improved over the course of the epidemic and could account for the decrease in complaints. These improvements are not specific to New York. There are no comparable studies from other major cities—if such studies were to show a similar downward trend it might be reasonable to attribute the decrease in 311 complaints in New York to improved pest management methods alone. Governmental and nongovernmental entities have also increased educational efforts, and these may have improved knowledge among landlords and residents (although this has not been formally assessed). Residents may choose to work directly with their landlord and not involve the city. If this were the case, we might expect to see areas in which mandated landlord reporting was out-of-step with 311 inquiries. Instead, we saw high concordance between the two (R^2^ = 0.60).

The decrease in bed bug complaints was observed across all New York City boroughs. However, the rate of decline was not equivalent. Higher income boroughs (Manhattan and Brooklyn) saw steeper declines than Queens and Staten Island. The differences in the rates of decline are very likely due to differences in financial means to properly treat infestations and incorporate recommendations of Integrated Pest Management (IPM). Differences in trust and access to city government and services are also likely to affect rates of decline^155,12,32–34^. In Chicago, bed bug infestation was highly associated with lower-income neighborhoods, crowding, and eviction notices. While we did not specifically assess sociodemographic features, it is likely that similar patterns exist in New York City and indeed have been noted in smaller scale surveys and assessments ^32,33^.

Despite substantial decreases in all boroughs, on the finer scale of NTD areas there are many persistently infested areas (Fig. 3). Many, though not all, of these persistently infested areas are located in lower-income areas. While New York City has made a vested effort to emphasize that tenants are not financially responsible for bed bug treatment, fear of eviction or cost (which are substantial) may prevent tenants from reporting bed bug infestation, which may promote spread to non-infested units. Areas with limited financial resources are therefore potentially at risk for persistent or entrenched bed bug infestation.

Our study was not without limitations. Self-reported and landlord reporting of bed bug infestation have inherent biases and inaccuracies ^33^. Sociodemographic differences have been documented between bed bug complaints and confirmed bed bug violations, with non-verified bed bug complaints (complaints that resulted in a negative bed bug inspection) occurring primarily in higher-income, majority white non-Hispanic neighborhoods ^33^. However, barring a standardized spatial sampling design, the data provided by New York City Open Data is likely the largest and most complete proxy available to estimate the spatial and longitudinal patterns in bed bug infestations.

Bed bugs are tied inextricably to their human hosts and the dynamic urban environment. Their resurgence in urban spaces and the necessity of rapid intervention strategies have elicited health policy, increased monitoring, and novel treatment strategies over the past 10 years. While there is continued need to increase active surveillance for bed bug infestations, particularly among vulnerable populations, the policy and public health approaches employed by New York City appear to be a step in the right direction.

## Data Availability

All data used for this project is available through NYC Open Data available from: https://opendata.cityofnewyork.us/

## Data Accessibility

All data is freely available from New York City Open Data. Data processing and code for tables and figures is available on Github.com/kphacker/NYC_bedbug_tables_figures

