## Supplemental for "Spatiotemporal Trends in Bed Bug Metrics, New York City"

**Supplementary material**

**Supplemental Table 1.** Descriptive properties of each of the datasets including timeframe, georeferencing information, organizational management of database, and codes used for analysis.

| Database | Timeframe Available | Georeferencing Information | Description | Code or text search used | Department |
|---|---|---|---|---|---|
| 311 Inquires | 2010–2019 | Not georeferenced | All calls, online inquiries, and inquiries registered through the app | N/A | NYC 311 |
| 311 bed bug specific requests | 2010–2019 | Not georeferenced | Official bed bug complaints, as well as general bed bug inquires | Text search using bed bug terms[2] | NYC 311 |
| Official bed bug complaints[3] | 2014–2019 | Georeferenced | Official bed bug complaints registered with HPD[4] | Problem Category Values: 2818 & 2517 | HPD |
| Official cockroach complaints | 2014–2019 | Georeferenced | Official cockroach complaints registered with HPD | Problem Category Values: 2514 & 2823 | HPD |
| Building owner reported bed bug infestations[5] | 2018 | Georeferenced | Units infested by bed bugs as reported by building managers to the HDP | Infested units | HPD |

[1] 311 bed bug specific requests are a subset of general 311 inquiries extracted through text search
[2] Text search terms included: bedbug(s), bed bug(s), and pest(s). Pests according to the housing maintenance code include bed bugs as well as "any unwanted member of class Insecta"(33)
[3] Formal bed bug complaints have been archived by the HDP since 2014 and are a subset of 311 inquiries
[4] Department of Housing and Preservation
[5] Report obtained by a freedom of information request

**Supplemental Table 2.** Total number of inquiries processed by 311 that included bed bugs as part of the description**.** Description of bed bug related inquires and the agencies that processed the request.

| Bed Bug Related Inquires | Departments Handling 311 Inquiries Regarding Bed Bugs (n = 246629) | | | | | | | |
|---|---|---|---|---|---|---|---|---|
| Inquiry Description | 3-1-1 Call Center | CUNY | DOE¶ | DOHMH# | HPD ‖ | HRA** | MTA†† | NYCHA‡‡ |
| BB* at CUNY§ College | 0 | 39 | 0 | 0 | 0 | 0 | 0 | 0 |
| Pests† in Apartment | 0 | 0 | 0 | 0 | 29479 | 0 | 0 | 0 |
| Pests† in Residential Public Area | 0 | 0 | 0 | 0 | 6479 | 0 | 0 | 0 |
| BB* complaint from a tenant in a residential building | 0 | 0 | 0 | 0 | 46886 | 0 | 0 | 0 |
| BB* complaint in business or nonprofit | 4423 | 0 | 0 | 0 | 0 | 0 | 0 | 0 |
| BB* complaint in domestic violence shelter | 0 | 0 | 0 | 0 | 0 | 270 | 0 | 0 |
| BB* complaint in hotel or SRO‡ | 0 | 0 | 0 | 0 | 1125 | 0 | 0 | 0 |
| BB* complaint in NYC school | 1 | 0 | 4596 | 0 | 0 | 0 | 0 | 0 |
| BB* complaint in NYC housing authority. | 2 | 0 | 0 | 0 | 0 | 0 | 0 | 12589 |
| Complaint that might attract pests† in subways or other MTA authority | 0 | 0 | 0 | 0 | 0 | 0 | 484 | 0 |
| BB* complaint in daycare centers | 12 | 0 | 0 | 418 | 0 | 0 | 0 | 0 |
| Get information about BB* in day care centers | 0 | 0 | 0 | 49 | 0 | 0 | 0 | 0 |
| Get information about how to comply with health commissioner's order about BB* | 3 | 0 | 0 | 2072 | 0 | 0 | 0 | 0 |
| Information for preventing and getting rid of BB* | 0 | 0 | 0 | 480 | 0 | 0 | 0 | 0 |
| Make a complaint about BB* in a residential building hotel or SRO‡ building | 0 | 0 | 0 | 0 | 106096 | 0 | 0 | 0 |
| Request a copy of BB* prevention and control brochure | 49 | 0 | 0 | 30939 | 0 | 0 | 0 | 0 |
| Information about filing BB annual report for building owners online | 0 | 0 | 0 | 0 | 138 | 0 | 0 | 0 |
| Total | 4484 | 39 | 4596 | 33958 | 190203 | 270 | 484 | 12589 |

*Bed Bugs, †Bed bugs, fleas, flies, roaches, mice, or other pests, ‡Single Room Occupancy, §City University of New York, ¶Department of Education, #Department of Health and Mental Hygiene, ‖Department of Housing Preservation and Development, **Human Resources Administration, ††Metropolitan Transit Authority, ‡‡New York City Housing Authority

**Supplemental Table 3.** Association between time and official bed bug and cockroach complaints accounting for seasonality. Model results of a linear harmonic model assessing the association between month and number of official bed bug complaints and cockroach complaints from 2014 – 2019. Bed bug and cockroach complaints were standardized by the total number of 311 inquiries to obtain percentages.

| Variable | Estimate | Confidence Interval | p-value |
|---|---|---|---|
| Bed Bug Complaints | | | |
| Time* | -1.8 e -5 | -2.4e-5– -1.2e-5 | $p < .01$ |
| Amplitude† | 0.3 | 0.1–0.5 | $p < .01$ |
| Phase Shift* | 1.4 | 0.8–2.0 | $p < .01$ |
| Cockroach Complaints | | | |
| Time* | 7.8e-5 | 6.5e-5–8.9e-5 | $p < .01$ |
| Amplitude† | 0.3 | 0.1–0.4 | $p < .01$ |
| Phase Shift* | 1.4 | 0.8–2.0 | $p < .01$ |

*Measured in months

†Measured in the number of complaints per total number of 311 inquires multiplied by 100,000

\

**Supplemental Table 4.** Bed bug complaints throughout the boroughs as processed by HPD by year.

| Borough | Year | Number of Bed Bug Complaints | Estimated Population | Proportion per 100,000 ± (95% Confidence Interval) |
|---|---|---|---|---|
| Bronx | Total | 10,394 | 1,434,093 | 724.8 ± CI (711, 738.8) |
| | 2014 | 1,330 | 1,413,566 | 94.09 ± CI (89.17, 99.28) |
| | 2015 | 2,337 | 1,428,357 | 163.6 ± CI (157.1, 170.4) |
| | 2016 | 1,993 | 1,436,785 | 138.7 ± CI (132.8, 144.9) |
| | 2017 | 1,968 | 1,455,846 | 135.2 ± CI (129.3, 141.3) |
| | 2018 | 1,561 | 1,437,872 | 108.6 ± CI (103.3, 114.1) |
| | 2019 | 1,205 | 1,432,132 | 84.14 ± CI (79.52, 89.03) |
| Brooklyn | Total | 14,544 | 2,598,602 | 559.7 ± CI (550.7, 568.8) |
| | 2014 | 2,009 | 2,570,801 | 78.15 ± CI (74.8, 81.64) |
| | 2015 | 3,300 | 2,595,259 | 127.2 ± CI (122.9, 131.6) |
| | 2016 | 2,925 | 2,606,852 | 112.2 ± CI (108.2, 116.3) |
| | 2017 | 2,589 | 2,635,121 | 98.25 ± CI (94.54, 102.1) |
| | 2018 | 2,172 | 2,600,747 | 83.51 ± CI (80.08, 87.1) |
| | 2019 | 1,549 | 2,582,830 | 59.97 ± CI (57.06, 63.03) |
| Manhattan | Total | 9,399 | 1,632,991 | 575.6 ± CI (564.1, 587.3) |
| | 2014 | 1,134 | 1,618,389 | 70.07 ± CI (66.11, 74.27) |
| | 2015 | 2,069 | 1,629,507 | 127 ± CI (121.6, 132.6) |
| | 2016 | 1,989 | 1,634,989 | 121.7 ± CI (116.4, 127.1) |
| | 2017 | 1,683 | 1,653,877 | 101.8 ± CI (97.02, 106.7) |
| | 2018 | 1,454 | 1,632,480 | 89.07 ± CI (84.61, 93.76) |
| | 2019 | 1,070 | 1,628,701 | 65.7 ± CI (61.88, 69.75) |
| Queens | Total | 6,485 | 2,301,409 | 281.8 ± CI (275, 288.7) |
| | 2014 | 812 | 2,280,602 | 35.6 ± CI (33.24, 38.14) |
| | 2015 | 1,374 | 2,301,139 | 59.71 ± CI (56.64, 62.95) |
| | 2016 | 1,303 | 2,310,011 | 56.41 ± CI (53.43, 59.55) |
| | 2017 | 1,118 | 2,339,280 | 47.79 ± CI (45.07, 50.68) |
| | 2018 | 1,032 | 2,298,513 | 44.9 ± CI (42.24, 47.72) |
| | 2019 | 846 | 2,278,906 | 37.12 ± CI (34.7, 39.71) |
| Staten Island | Total | 907 | 473,926 | 191.4 ± CI (179.3, 204.2) |
| | 2014 | 145 | 471,522 | 30.75 ± CI (26.13, 36.19) |
| | 2015 | 166 | 472,481 | 35.13 ± CI (30.18, 40.9) |
| | 2016 | 187 | 473,324 | 39.51 ± CI (34.23, 45.59) |
| | 2017 | 157 | 475,948 | 32.99 ± CI (28.21, 38.57) |
| | 2018 | 163 | 474,101 | 34.38 ± CI (29.49, 40.08) |
| | 2019 | 89 | 476,179 | 18.69 ± CI (15.18, 23.01) |

**Supplemental Figures**

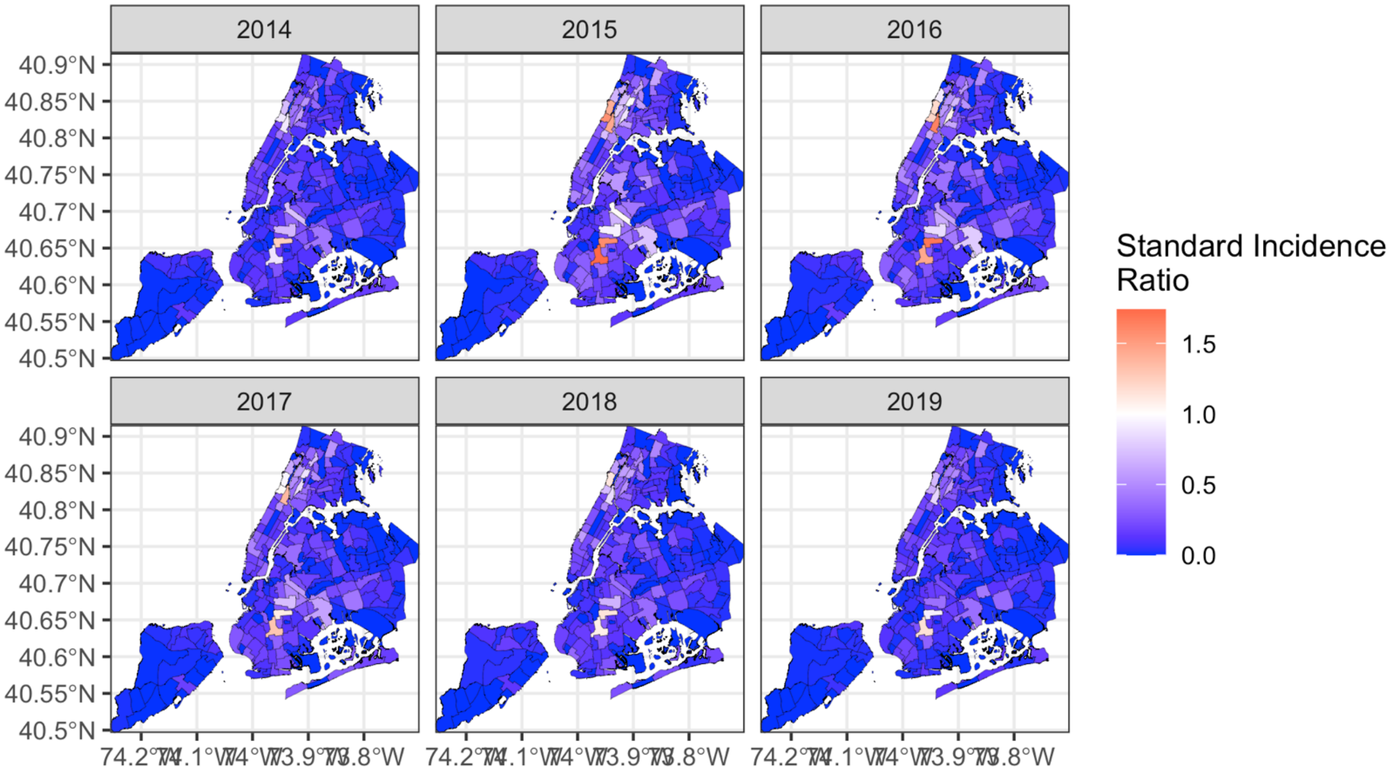


**Supplementary Fig. 1.** Standard Incident Ratio of official bed bug complaints per NTA area from 2014–2019.

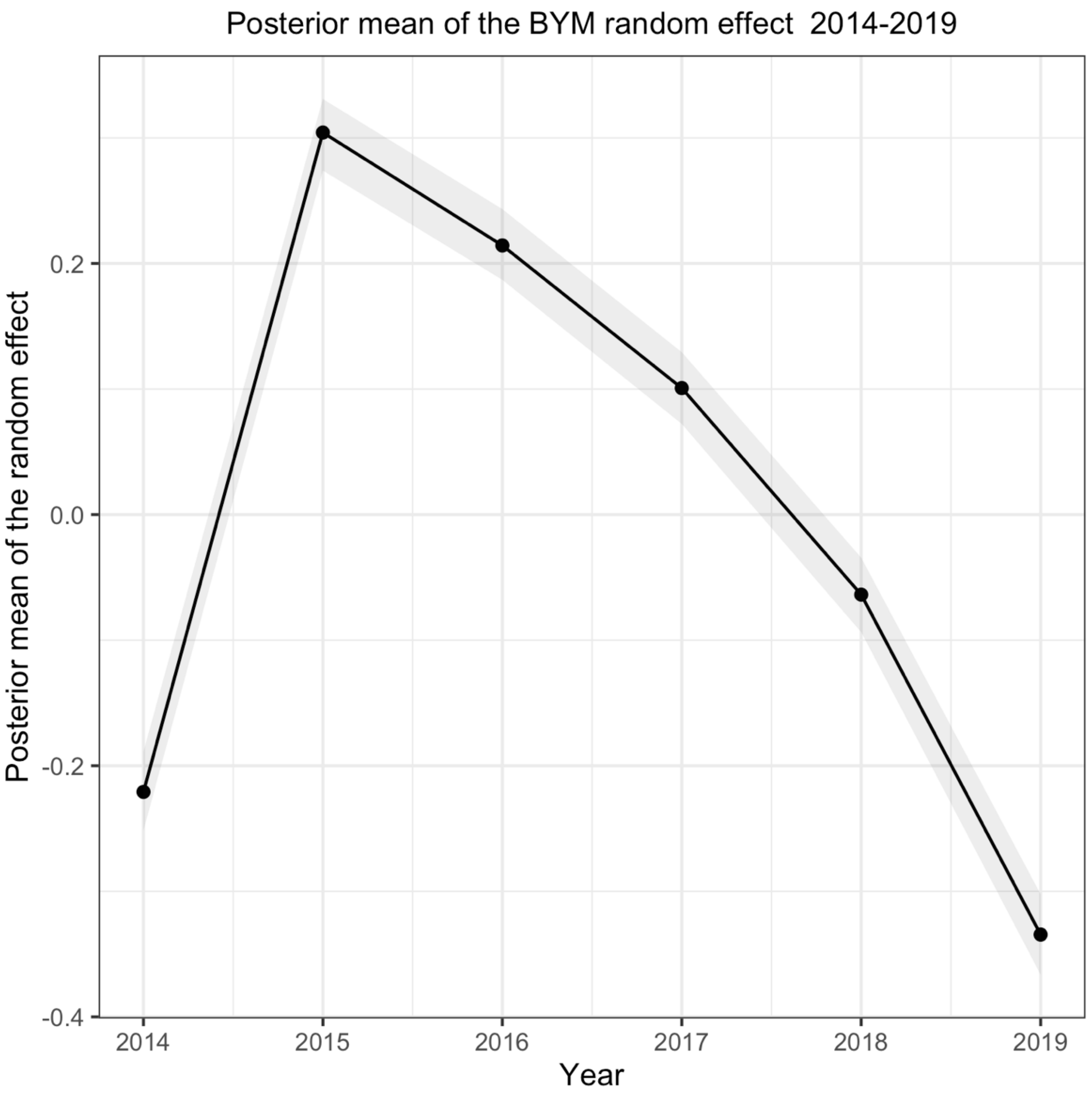


**Supplementary Fig. 2.** Plotted posterior mean of the BYM random effect of the from 2014–2019.

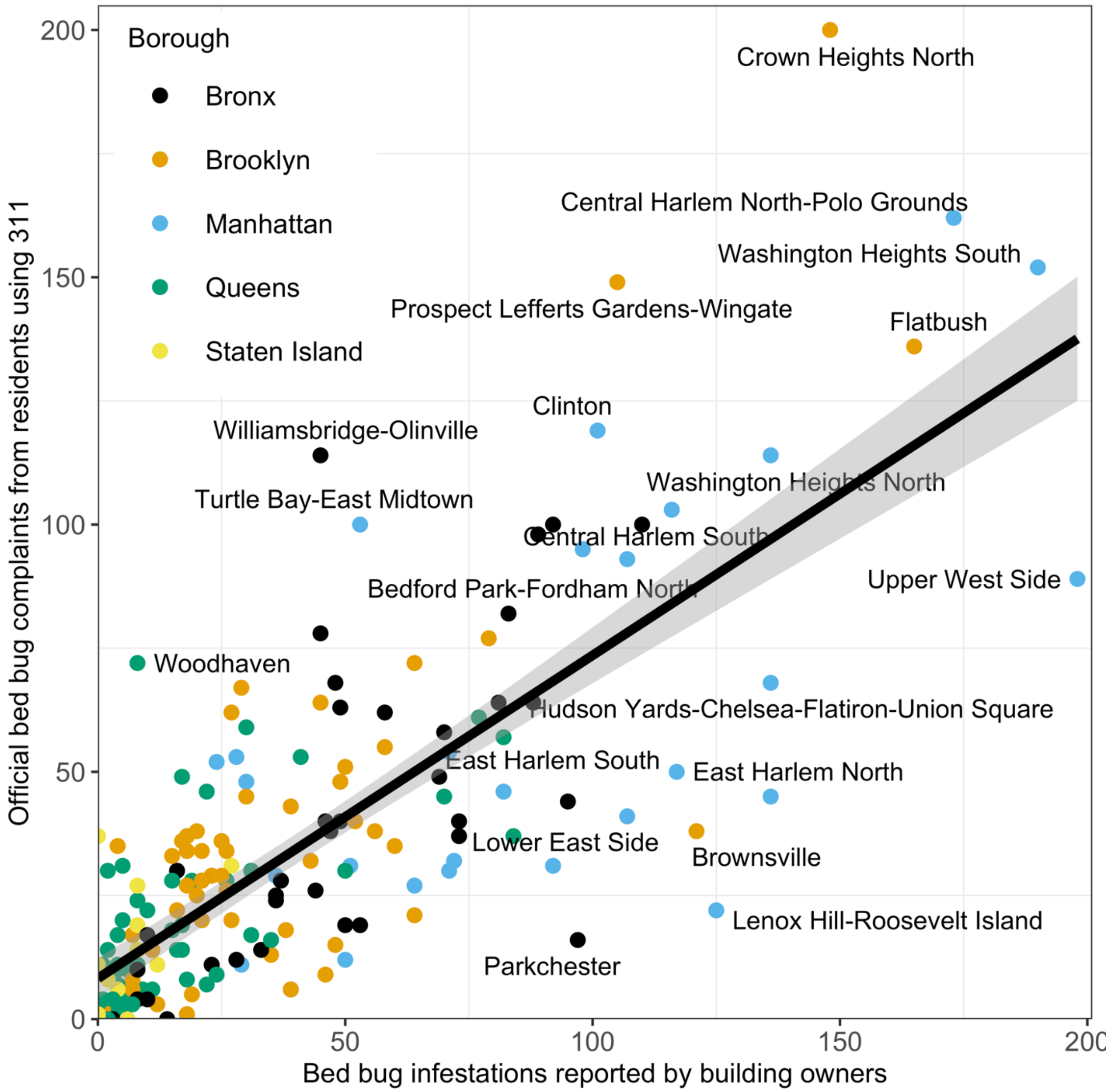


**Supplementary Fig. 3.** Comparison between building manager reported bed bug infestation and official bed bug complaints registered to HPD for 2018. Correlation between official bed bug complaints from residents and building manager reported infestation was high ($R^2 = 0.60$).

**Supplemental Citations:**


New York City Laws, Maintenance Codes, Resident Information, and 311 information

External Datasets Used

*New York City Community Health Survey*

7. NYC Health. Community Health Survey [Internet]. 2020 [cited 2020 Apr 26]. Available from: https://www1.nyc.gov/site/doh/data/data-sets/community-health-survey.page

*New York City 311 Call Center*

8. City of New York Open Data. 311 Call Center Inquiry. City Government. 13 June 2016. Available from: https://data.cityofnewyork.us/City-Government/311-Call-Center-Inquiry/wewp-mm3p.

*New York City Bed Bug Complaints*

9. City of New York Open Data. Housing Maintenance Code Complaints. Housing and Development. 2020 [cited: 10 March 2020]. Available from: https://data.cityofnewyork.us/Housing-Development/Housing-Maintenance-Code-Complaints/uwyv-629c

*New York City Bed Bug Complaints Georeferenced*

10. City of New York Open Data. Complaint Problems. Housing and Development. 2020 [cited: 10 March 2020]. Available from: https://data.cityofnewyork.us/Housing-Development/Complaint-Problems/a2nx-4u46

Software Used

11. *QGIS Geographic Information System.* Open Source Geospatial Foundation Project http://qgis.org

12. R: A language and environment for statistical computing [computing software]. Version 1.3.1093. Vienna, Austria; 2017.

Bed bugs in the media

13. BedBug Central. History and Resurgence [internet]. 2008 [cited 2020 Apr 26]. Available from: https://www.bedbugcentral.com/bedbugs101/history-resurgence

14. Buckley C. Legislature Passes Bedbug-Notification Law. The New York Times [Internet]. 2010 Jun 24 [cited: 26 Apr 2020]; Available from: https://www.nytimes.com/2010/06/25/nyregion/25bedbugs.html

15. Pilkington E. How bedbugs invaded New York. *The Guardian*. https://www.theguardian.com/world/2010/oct/21/bedbugs-invaded-new-york. Published October 21, 2010. Accessed April 26, 2020.
